# The effects of daylight saving time clock changes on accelerometer-measured sleep duration in the UK Biobank

**DOI:** 10.1101/2024.05.28.24308011

**Authors:** Melanie A de Lange, Rebecca C Richmond, Kate Birnie, Chin Yang Shapland, Kate Tilling, Neil M Davies

## Abstract

We explored the effects of daylight saving time (DST) clock changes on sleep duration in a large accelerometer dataset. Our sample included UK Biobank participants (n= 11,780; aged 43-78) with accelerometer data for one or more days during the two weeks surrounding the Spring and Autumn DST transitions from October 2013 and November 2015. Between-individual t-tests compared sleep duration on the Sunday (midnight to midnight) of the clock changes to the Sunday before and the Sunday after. We also compared sleep duration on all other days (Monday-Saturday) before and after the clock changes. In Spring, mean sleep duration was 65 minutes lower on the Sunday of the clock changes than the Sunday before (95%CI -72 to -58 minutes) and 61 minutes lower than the Sunday after (95%CI -69 to -53). In Autumn, the mean sleep duration on the Sunday of the clock changes was 33 minutes higher than the Sunday before (95% CI 27 to 39 minutes) and 38 minutes higher than the Sunday after (95% CI 32 to 43 minutes). There was some evidence of catch-up sleep after both transitions, with sleep duration a little higher on the Monday - Friday than before, although this was less pronounced in Autumn. Future research should use large datasets with longer periods of accelerometer wear to capture sleep duration before and after the transition in the same individuals, and examine other aspects of sleep such as circadian misalignment, sleep fragmentation or daytime napping.

## 1. INTRODUCTION

Daylight saving time (DST), the practice of moving the clocks one hour forward in Spring and one hour back in Autumn, was introduced in World War One to maximise exposure to daylight during the working day and reduce energy use^1^. It operates in around 70 countries, including the UK, and affects over a quarter of the world’s population^2, 3^. However, growing evidence suggests that the DST clock changes may adversely affect population health. For example, studies have found an increased incidence of myocardial infarction^4^, atrial fibrillation^5^, strokes^6^, and fatal traffic accidents^7^ in the weeks immediately following the Spring clock change, as well as an increased risk of depressive episodes after the Autumn transition^2^. These health consequences are thought to be mediated by sleep deprivation and circadian disruption^2, 6–8^.

Previous research has reported that both Spring and Autumn clock changes were associated with less sleep for around a week after the change^9^. Sleep after the Spring change was found to be more fragmented, and people took longer to get to sleep, whilst sleep loss after the Autumn change was due to people continuing to wake early^9^. However, estimates of the exact effects of clock changes on sleep duration vary. Studies report that sleep duration on the night of the Spring clock change was 5 - 30 minutes less than on the same night on surrounding weekends^10–12^, whilst estimates of sleep duration on the night of the Autumn clock change range from no difference to 40 minutes more than the surrounding weekends^11–14^. Some studies report that sleep duration on the weekdays after the Spring changes is up to 60 minutes less than surrounding weeks, whilst others report that it is up to 28 minutes more^11, 14–19^. Estimates of sleep duration on the weekdays after the Autumn changes are also conflicting, with some studies estimating that it is up to 25 minutes less than surrounding weeks, whilst other studies report it being up to 11 minutes more^11, 14, 15, 17, 20^.

Potential modifiers of the effect of the clock changes on sleep duration include chronotype (morning/evening preference) and habitual sleep duration. The Spring change is thought to have the most detrimental effect on the sleep duration of evening types and short sleepers (<8 hours), whilst morning types and long sleepers (>8 hours) lose the most sleep in Autumn^21–25^. Studies also indicate that those not meeting their preferred sleep duration the week before the clock change are most disrupted by both changes^16^.

Many studies of the effects of DST transitions on sleep have relied on subjective self-report data^10, 11, 14, 15, 19, 20^, which may be affected by recall bias and have been shown to only moderately correspond to more objective measures of sleep, such as actigraphy^26^. Whilst some DST research has used actigraphy data, most of these studies have suffered from small sample sizes (n < 100)^13, 16–18^, reducing their estimates’ precision. In this study, we used accelerometer data from the UK Biobank to explore the effects of the DST clock changes on sleep duration in a much larger sample (n= 11,780).

## 2. METHODS

### 2.1 Study Population

The UK Biobank is a prospective cohort study with data on around 500,000 people aged 40-69 years recruited from across the UK in 2006-2010^27^ (5.5% participation rate^28^). At baseline, participants completed questionnaires covering sociodemographic and lifestyle factors and their medical history. Participants also had physical (e.g. blood pressure) and biological (e.g. blood/urine samples) measures taken^27^. 236,519 participants who had provided an email address were randomly selected and invited to wear an accelerometer for seven days between June 2013 and December 2015. Of these, 106,053 agreed (participation rate 44.8%)^29^ , and 103,712 ultimately provided accelerometer data. After subsequent participant withdrawals, accelerometer data was available for 103,628 participants (See Figure 1).

**Figure 1.**
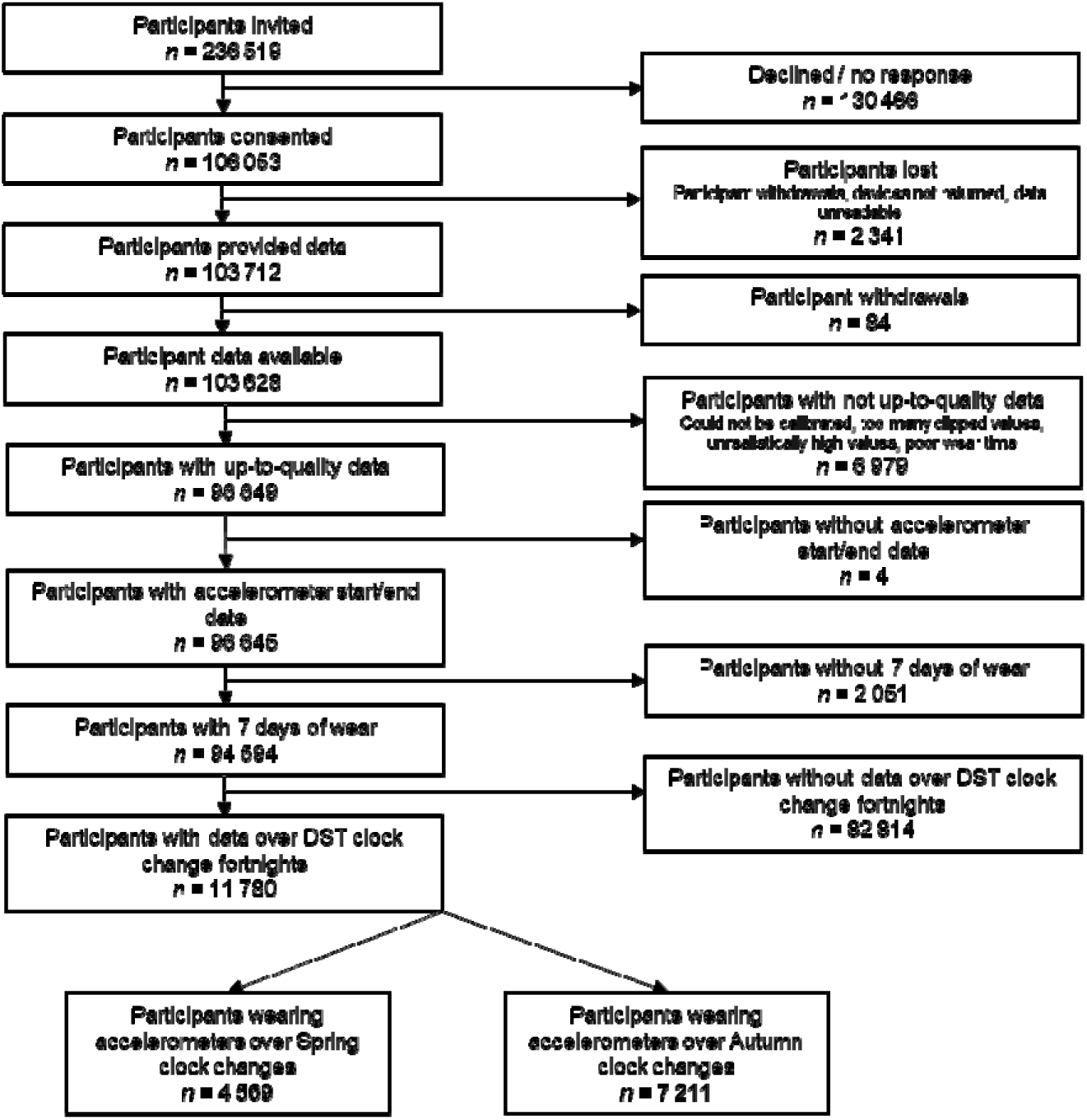
Participant flow diagram

In keeping with previous analyses^29, 30^, for quality control, we excluded participants if they had poor wear time (failing to have ≥ three days of data and data in each hour of the 24-hour day over multiple days), poor calibration (insufficient data preventing recalibration by the previous or next measurement), unrealistically high activity levels (an average overall volume of activity per day of over 100 milli-gravity units (m*g*)), or if over 1% of values were clipped before or after calibration (a clipped value is when the measured value exceeds the maximum or minimum values the device can measure and therefore any excess acceleration is not recorded). We also excluded records of those without accelerometer wear start and end dates and those with less than seven days of wear.

In this study, participants wore Axivity AX3 triaxial accelerometers on their dominant wrist (described in detail elsewhere^29^). Participants were asked to wear the accelerometer continuously and complete their daily activities as normal^29^. They wore the accelerometers for 168 hours (7 x 24 hours), starting at 10am on day one and finishing at 10am on day eight. Consequently, day one data was made up of data from 10am to midnight on day one, combined with data from midnight to 10am on day eight. For our study, this meant we had to exclude day one for all participants as it consisted of data from more than one day. We therefore only included participants who had data (excluding their day one) for any of the days during the two weeks surrounding the DST clock changes (the 15 days from the Sunday before the clock change to the Sunday after). This resulted in a final sample of 11,780 participants, 4,569 in the Spring and 7,211 in the Autumn (See Figure 1). Our study covered two Spring clock changes (2014 and 2015) and three Autumn clock changes (2013, 2014 and 2015) .

UK Biobank participants started wearing their accelerometers on different days of the week, with no participants starting on a Tuesday or Sunday. In our final sample, participants started to wear their accelerometer from two Mondays before the clock changes to the Saturday after the clock changes. Having excluded data from the first day for each participant, our resulting dataset included data from the Sunday before the clock changed to the Sunday after the clock changed, with different numbers of participants per day (See Table S1). No participants had data for the whole study period.

### 2.2 Sleep duration

Sleep duration has been derived from the raw UK Biobank accelerometer data by Doherty et al. (2017) ^29^. This includes the proportion of time (from midnight to midnight) spent asleep overall, by day, on weekends and weekdays, and broken down by hour (overall, on weekends and weekdays). Doherty et al. adjusted the data of those wearing the accelerometers over the clock changes so that the Sundays of the change comprised 24 hours (instead of 23 or 25). In Spring (when the clocks go forward an hour at 1 am and we lose an hour), they imputed an hour of data for the hour missing between 1 and 2 am. In Autumn (when the clocks go back an hour at 2 am and we gain an hour), they overwrote the data for the hour between 1 and 2 am (effectively deleting an hour of data).

To accurately capture the effects of the clock changes in this study, we adjusted the sleep proportion on the Sunday of the clock changes to add/subtract the hour deleted/imputed by Doherty et al. The sleep proportion for the hour that we added/subtracted was the average sleep proportion between 1-2 am for the 96,645 UK Biobank participants who wore accelerometers on normal weekends (0.90). In Spring, our sleep proportion on the Sunday of the clock changes was calculated by the following formula: (Doherty sleep proportion on Sunday x 24 - 0.90) / 23. In Autumn, the following formula was applied: (Doherty sleep proportion on Sunday x 24 + 0.90) / 25. We then rescaled the sleep proportion data to duration in minutes by multiplying data for Monday-Saturday and normal Sundays by 1440 (the number of minutes in 24 hours). Data for the Sundays of the clock changes was multiplied by 1,380 in Spring (to adjust for the Sunday only having 23 hours) and by 1,500 in Autumn (to adjust for the Sunday having 25 hours). For examples of these adjustments, see Data S1.

### 2.3 Sociodemographic characteristics

Several sociodemographic characteristics were considered potential effect modifiers of the effect of the clock changes on sleep duration. Sex (male or female) was obtained from a central registry at recruitment, but sometimes updated by the participant. Age (43-58, 59-67 or 68-78) was calculated using age at recruitment and date of accelerometer wear. Other potential effect modifiers were self-reported current employment status (employed, retired or other), self-reported habitual sleep duration in whole hours (<=6 hours, 7-8 hours or >=9 hours) and self-reported chronotype (morning, no preference or evening). Detailed information on how the sociodemographic variables were derived is provided in Data S1.

### 2.4 Statistical Analysis

Mean daily sleep duration (from midnight to midnight) for the two weeks surrounding the Spring and Autumn clock changes was plotted (see Table S1 for sample sizes per day). We conducted between-individual t-tests to compare participants’ sleep duration on the Sunday of the clock changes to the Sunday before and the Sunday after the transitions. We also conducted between-individual t-tests to compare sleep duration on all other days (Monday-Saturday) before and after the clock changes. To establish whether sociodemographic factors modified the effects of the clock changes, all analyses were then stratified separately by age, sex, current employment status, self-reported habitual sleep duration, and chronotype. Cochran’s Q was used to test heterogeneity between strata. It was impossible to conduct within-person comparisons of sleep duration on the Sunday of the clock change to the Sunday before and after, or the same weekdays before and after, because participants only had six days of usable accelerometer data each.

Analyses were performed in Stata version 16 via JupyterLab in DNA Nexus. The full code for data cleaning and analysis is available at https://github.com/MeldeLange/dst_accel. This research was pre-specified as part of UK Biobank application 86626.

## 3. RESULTS

### 3.1 Participant characteristics

In this study, 44% of participants were male, 66% were aged 59 and over, and 32% were retired (see Table 1). In our sample, 72% reported sleeping for 7-8 hours per night, and 57% stated they were a morning chronotype.

**Table 1.** Sample characteristics.

|  | **Total Sample** | **Spring clock change** | **Autumn clock change** |
| --- | --- | --- | --- |
| **Total (n)** | 11,780 | 4,569 | 7,211 |
| **Variables n (%)** |  |  |  |
| Sex |  |  |  |
| Male | 5,165 (43.8%) | 1,975 (43.2%) | 3,190 (44.2%) |
| Age |  |  |  |
| Tertile 1 (age 43-58) | 3,988 (33.9%) | 1,599 (35.0%) | 2,389 (33.1%) |
| Tertile 2 (age 59-67) | 4,486 (38.1%) | 1,707 (37.4%) | 2,779 (38.5%) |
| Tertile 3 (age 68-78) | 3,306 (28.1%) | 1,263 (27.6%) | 2,043 (28.3%) |
| Current employment status |  |  |  |
| Paid employment/self-employed | 7,141 (60.8%) | 2,815 (61.8%) | 4,326 (60.1%) |
| Retired | 3,769 (32.1%) | 1,426 (31.3%) | 2,343 (32.6%) |
| Other | 842 (7.2%) | 313 (6.9%) | 529 (7.3%) |
| Sleep duration (hours) (self-reported) |  |  |  |
| <=6 hours | 2,519 (21.5%) | 999 (21.9%) | 1,520 (21.2%) |
| 7-8 hours | 8,467 (72.2%) | 3,270 (71.8%) | 5,197 (72.4%) |
| 9 or more hours | 748 (6.4%) | 284 (6.2%) | 464 (6.5%) |
| Chronotype (self-reported) |  |  |  |
| Morning | 6,707 (57.4%) | 2,558 (56.4%) | 4,149 (58.0%) |
| No preference | 1,209 (10.3%) | 473 (10.4%) | 736 (10.3%) |
| Evening | 3,777 (32.3%) | 1,507 (33.2%) | 2,270 (31.7%) |

### 3.2 Mean sleep duration by day

Figures 2 and 3 show mean sleep duration by day over the fortnight surrounding the Spring and Autumn clock changes (see Table S1 for sample sizes per day and Table S2 and Figures S1-10 for stratified results). Sleep duration on the Sunday of the clock changes was lower (in Spring) and higher (in Autumn) than on all other days, including the previous and subsequent Sundays. In general, sleep duration tended to be a little higher on the Monday - Friday after both clock changes than before, although this difference was less pronounced in Autumn.

**Figure 2.**
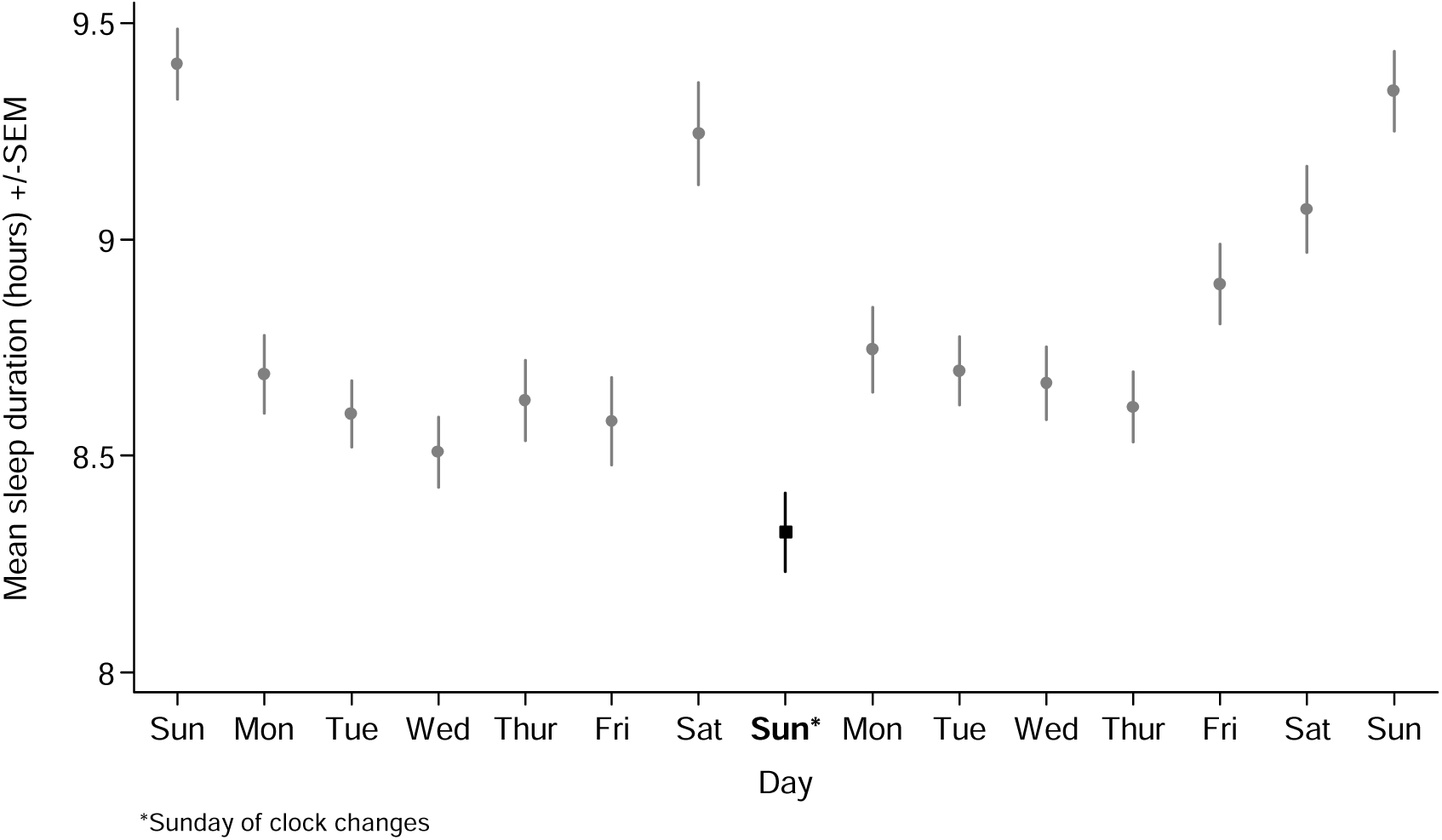
Mean daily sleep duration by day over the Spring clock changes. Dot/square shows the mean. Bar shows the standard error of the mean.

**Figure 3.**
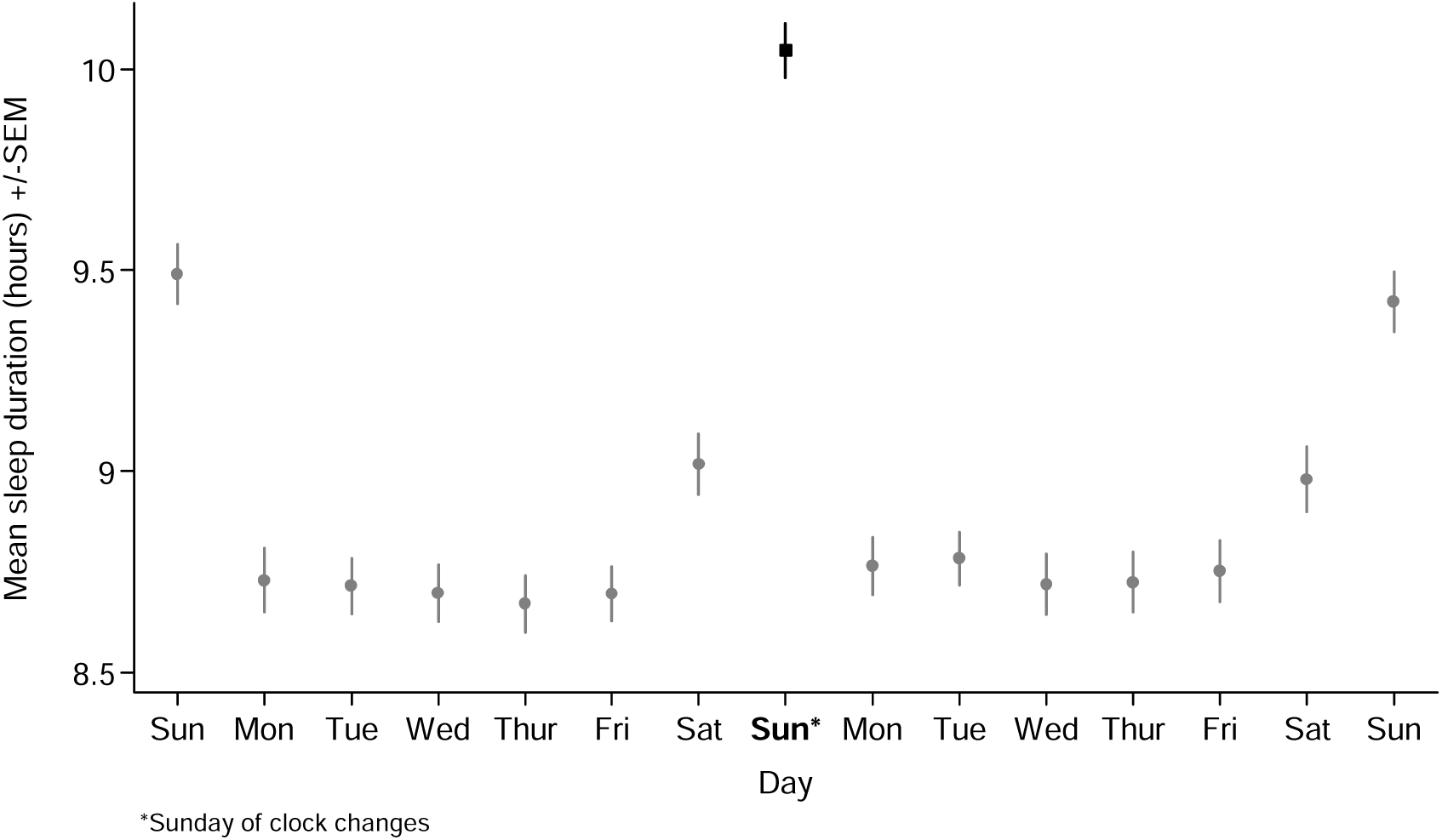
Mean daily sleep duration by day over the Autumn clock changes. Dot/square shows the mean. Bar shows the standard error of the mean.

### 3.3 Between-individual comparisons of sleep duration on the Sunday of the clock change versus the Sunday before and the Sunday after

Between-individual t-tests comparing people’s sleep duration on the Sunday of the clock changes to the Sunday before and the Sunday after are shown in Table 2 (see Table S3 for stratified results). In Spring, mean sleep duration on the Sunday of the clock changes (8 hours 19 minutes) was 65 minutes lower than the Sunday before (95% CI -72 to -58 minutes) and 61 minutes lower than the Sunday after (95% CI -69 to -53).

**Table 2.** Between-individual comparisons of mean sleep duration (hours and minutes) on the Sunday the clock changes vs the Sunday before and the Sunday after.

|  | Sun clock change |  | Sunday before |  |  |  | Sunday after |  |  |  |
| --- | --- | --- | --- | --- | --- | --- | --- | --- | --- | --- |
|  | n | Mean sleep duration (SD) | n | Mean sleep duration (SD) | Difference in minutes (SE) | p value* | n | Mean sleep duration (SD) | Difference in minutes (SE) | p value* |
| Spring | 1515 | 8 hrs 19 mins<br>(1 hr 48 mins) | 1785 | 9 hrs 24 mins<br>(1 hr 45 mins) | -64.87 (3.72) | $3.667 \times 10^{-65}$ | 1269 | 9 hrs 21 mins<br>(1 hr 41 mins) | -61.14 (4.00) | $9.902 \times 10^{-51}$ |
| Autumn | 2688 | 10 hrs 3 mins<br>(1 hr 47 mins) | 2277 | 9 hrs 29 mins<br>(1 hr 46 mins) | 33.39 (3.04) | $8.065 \times 10^{-28}$ | 2246 | 9 hrs 25 mins<br>(1 hr 46 mins) | 37.51 (3.05) | $2.677 \times 10^{-34}$ |

SD: Standard deviation. SE: Standard error. \*P value from t-tests.

In Autumn, mean sleep duration on the Sunday of the clock changes (10 hours 3 minutes) was 33 minutes higher than the Sunday before (95% CI 27 to 39 minutes) and 38 minutes higher than the Sunday after (95% CI 32 to 43 minutes). There was little evidence of differences by sociodemographic characteristics for either transition, with lower sleep on the Sunday of the Spring transition and higher sleep on the Sunday of the Autumn transition seen for all sociodemographic subgroups (see Table S3 for stratified results and Cochran’s Q test of heterogeneity results).

### 3.4 Between-individual comparisons of sleep duration on Monday to Saturday after the clock change versus the same day the week before

Table 3 highlights in greater detail the trend over both Spring and Autumn transitions of sleep duration being higher on the Monday to Friday after the clock changes than before, as well as being lower on the Saturday after the clock change than the Saturday before. However, as already mentioned, this pattern was much less marked in Autumn. The mean difference in sleep duration between weekdays (Monday-Friday) after and before the Spring clock change was 7.4 minutes, whereas in Autumn the average weekday difference was 2.9 minutes. We detected differences by sex, with women generally tending to sleep for fewer minutes on weekdays after the Spring clock change than before, whilst men generally slept for longer on weekdays after the Spring clock change than before. This pattern (albeit on a smaller scale) was also seen after the Autumn change. There was no obvious pattern to the changes after the Spring clock changes, across age groups. After the Autumn clock change, a general pattern was that the youngest tertile exhibited a positive change (slept longer after the clock change), the middle tertile had a smaller positive or negative change, and the oldest tertile tended to have a negative change (See Figures S1-5 and Table S4 for stratified results and Cochran’s Q test of heterogeneity).

**Table 3.** Between-individual comparisons of mean sleep duration (hours and minutes) on the Monday to Saturday before and after the Spring and Autumn clock changes.

|  | Spring |  |  |  | Autumn |  |  |  |
| --- | --- | --- | --- | --- | --- | --- | --- | --- |
|  | n | Difference in sleep duration (minutes) | 95% CI | p value | n | Difference in sleep duration (minutes) | 95% CI | p value |
| Mon after vs before | 2,518 | 3.45 | -4.52,11.42 | 0.396 | 3,839 | 2.17 | -4.26,8.61 | 0.508 |
| Tue after vs before | 3,646 | 5.93 | -0.65,12.50 | 0.077 | 4,935 | 4.12 | -1.57,9.81 | 0.156 |
| Wed after vs before | 3,018 | 9.53 | 2.51,16.56 | 0.008 | 4,080 | 1.36 | -4.81,7.53 | 0.666 |
| Thur after vs before | 2,906 | -0.95 | -8.37,6.48 | 0.803 | 3,945 | 3.26 | -2.94,9.46 | 0.303 |
| Fri after vs before | 2,491 | 19.01 | 10.80,27.22 | $5.945 \times 10^{-06}$ | 4,309 | 3.38 | -2.68,9.45 | 0.274 |
| Sat after vs before | 2,043 | -10.46 | -19.76,-1.16 | 0.028 | 3,857 | -2.28 | -8.94,4.38 | 0.502 |

## 4. DISCUSSION

In this study, we used accelerometer data from 11,780 UK Biobank participants to examine the effects of the DST clock changes on sleep duration. In the Spring, our data showed that the mean sleep duration was 65 minutes lower on the Sunday of the clock changes than on the Sunday before and 61 minutes lower than on the Sunday after. These results suggest that the Spring transition is associated with reduced sleep duration. It is difficult to directly compare our results which look at sleep duration each day (from midnight to midnight), to studies that examine sleep each night (from noon to noon)^10, 12^. However, we found a loss of sleep greater than that seen in previous studies which have reported a 5-30 minute reduction in sleep duration on the night of the Spring clock change^10, 12^ and a study reporting a 30 minute reduction on the Sunday of the change^11^.

In Autumn, we found that mean sleep duration was 33 minutes higher on the Sunday of the clock changes than the Sunday before and 38 minutes higher than on the Sunday after. This suggests that the Autumn transition is associated with increased sleep duration. However, participants did not (or could not) take advantage of the full extra hour of increased sleep. This may be because their body clocks or social commitments prevent them from having an entire hour more sleep. Our results are consistent with the existing literature, with one earlier study reporting a 20-minute increase on the night of the Autumn clock change compared to surrounding weekends^12^ and another reporting a 40-minute increase in sleep on the Sunday of the change^11^.

We found that sleep duration was higher on the weekdays after both the Spring and Autumn clock changes, although this trend was more pronounced in Spring. Prior studies of the Spring transition have reported sleep duration on the following weekdays being up to 60 minutes lower^14–16, 18, 19^, no different^11^ or up to 28 minutes higher^17^ than the previous week. Likewise, studies have reported weekday sleep duration after the Autumn change being 25 minutes lower^17^, no different^11, 14, 15^ or 11 minutes more^20^ than the previous week. Our estimates of weekday differences therefore fall within the range reported by previous studies. The fact that we find some weak evidence of catch up sleep after the Autumn clock change (when we gain an hour’s sleep) suggests that the disruption to circadian rhythms caused by the clock change (rather than just lost sleep on the night of the clock change) could potentially be leading people to be more tired and sleep for longer.

Some research suggests that catching up on sleep can help people to recover from the effects of sleep loss on pain tolerance, mood, fatigue, cognitive performance and metabolic function^31–34^. However, other studies have found that recovery sleep is insufficient to fully restore brain or metabolic function^35–37^. How long it takes to recover from acute sleep loss is not entirely clear, with one study reporting it can take up to four days to recover from one hour of lost sleep^38^ and another indicating that three nights of sleep are needed to make up for one night of insufficient sleep^39^. Future studies are needed to shed light on whether recovery sleep after the Spring clock change benefits health and, if so, how much recovery sleep is needed.

Overall, our data indicate that the negative effects of the clock changes on sleep duration in Spring are more short-lived than some previous studies suggest. Most importantly, we did not find evidence that the sleep loss associated with the Spring transition lasted for a whole week. That said, the acute sleep loss observed in this study is not necessarily inconsequential for health, as research suggests that just one night of sleep loss (<6 hours of sleep) is associated with an immediate decline in self-reported mental health, as well as an increase in the number and severity of physical health symptoms^40^. Furthermore, the clock changes themselves have been associated with an increase in cardiovascular events^4–6^, traffic accidents^7^ and depressive episodes^2^.

A strength of this study is the large sample size, aided by data from multiple years, which meant our overall estimates were more precise than those of previous studies. In addition, we had data for both Spring and Autumn clock changes, which enabled us to compare the effects of the transitions. Furthermore, this study benefited from objectively assessed sleep duration using wrist-worn accelerometers. When tested, the machine learning model used to create the sleep data in this study correctly identified sleep with a very high level of accuracy^30^.

That being said, our study has several limitations. Unlike previous studies, participants did not have data for the whole study period. As a result, within-individual comparisons were not possible. Secondly, the UK Biobank does not represent the wider UK population, with participants being older, more affluent and healthier than non-participants^28^. In addition, self-reported lifestyle factors such as sleep duration, chronotype and employment status were collected at baseline so may not have been accurate when the participants wore the accelerometers several years later. Finally, our dataset only included information on sleep duration, not the timing of sleep, so it was outside the scope of the current study to explore the effects of the clock changes on circadian rhythms.

In conclusion, this study adds to the growing body of evidence that the shift forward to daylight saving time in Spring is associated with an acute loss of sleep. However, we found that this was more short-lived than previous, smaller studies suggest. While sleep loss occurred on the Sunday of the Spring clock change, sleep duration on the weekdays following the transitions was not adversely affected. In fact, there was some evidence of catch up sleep after both clock changes. Future research should use large datasets with longer periods of accelerometer wear to capture sleep duration before and after the transition in the same individuals. It could also examine other aspects of sleep architecture such as circadian misalignment, sleep fragmentation, daytime napping/dozing or time asleep versus time awake.

## Author contributions

N.M.D and M.A.dL developed the study concept. M.A.dL performed the data analysis and drafted the first version of the manuscript. M.A.dL, N.M.D, R.C.R, K.T, K.B and C.Y.S interpreted the data, reviewed and revised the manuscript, and approved the final version as submitted.

## Supporting information

Data_S1_SuppInfo

Figures_S1-S10_SuppInfo

Tables_S1-S4_SuppInfo

## Data availability statement

The UK Biobank dataset used to conduct the research in this paper is available via application directly to the UK Biobank. Applications are assessed to meet the required criteria for access, including legal and ethical standards. More information regarding data access can be found at https://www.ukbiobank.ac.uk/enable-your-research. Full code for all analyses is available at https://github.com/MeldeLange/dst_accel

