## Supplementary material for "The effects of daylight saving time clock changes on accelerometer-measured sleep duration in the UK Biobank": Data_S1_SuppInfo

**Data S1: Supplementary method**

**1) Sociodemographics**

Data on sociodemographics was self-reported by participants via a touchscreen questionnaire completed at their initial UK Biobank assessment centre visit (Questionnaire available at https:/ /biobank.ctsu.ox.ac.uk/showcase/ukb/docs/TouchscreenQuestionsMainFinal.pdf).

We collapsed discrete variables to make ordinal variables in order to facilitate stratifying our analyses and recoded categorical variables to reduce the number of categories.

**Sex**

Sex (male/female) was taken from the central NHS registry at recruitment, but was updated by participants in some cases.

**Age**

Participants’ age when they started to wear the accelerometer was derived by calculating the difference (in whole years) between the date when participants started wearing the accelerometer and the date they attended the baseline assessment centre. This difference was then added to the age of participants at baseline (whole years). We collapsed this variable into three age tertiles: 43-59, 60-67, 68-77.

**Current employment status**

We reduced current employment status from 9 to 3 categories. Participants could choose more than one answer. Anyone that chose “employed” or “self-employed” in any answer was recoded as “Employed”, anyone who chose “Retired” in any answer was recoded as “Retired”. Participants not falling in these two categories (including "None of the above") were included in a final ‘Other’ category. Those who chose “prefer not to answer” were coded as missing.

**Sleep duration**

Self-reported sleep duration was derived from the question "About how many hours sleep do you get in every 24 hours? (please include naps)". Participants could only answer in whole hours. We recoded those choosing “Prefer not to answer” or “Do not know” as missing. We recoded extreme values of sleep duration (over 18 hours and under three hours) to missing. We then collapsed the discrete variable into three categories “<=6 hours”, “7-8 hours” and “>=9 hours”.

**Chronotype**

Self-reported chronotype was derived from participants’ answers to whether they considered themselves to be: “Definitely a morning person”, “More a morning than evening person”, “More an evening than a morning person”, “Definitely an evening person”, “Do not know” or “Prefer not to answer”. We coded “Prefer not to answer” as missing and “Do not know” as “No preference”. From this we derived a three point ordinal variable for chronotype: “Morning”, “No Preference” and “Evening”.

**2)** **Example of adjusting the Sunday of the clock changes from a 24 hour day to 23 or 25 hours**

**a) Assuming someone who sleeps for 8 hours between 10pm and 6am.**

**Spring**

The clocks go forward an hour at 1am.

On the Saturday you sleep from 10pm to midnight.

On the Sunday you sleep from midnight to 6am (but it is now 7am). You then sleep from 10pm to midnight on the Sunday evening.

The hour from 1-2am does not exist in the accelerometer records, but it records you waking up at 7am instead of 6am so it still records you sleeping for 8 hours on the Sunday:

12-1am, 2-3am, 3-4am, 4-5am, 5-6am, 6-7am, 10-11pm,11-12pm

But this is out of a 23 hour day so sleep proportion = 8/23 = 0.347826.

To turn it from a 23 hour day to a 24 hour day Doherty et al. imputed sleep for the hour 1-2am and added this in. e.g. (0.347826 x 23 +0.9) / 24 = 0.370833.

Doherty et al used data from other days at the same time day of day to create the imputed value.

Here we’ve calculated 0.9 as a comparative sleep proportion for the hour that Doherty added. 0.9 is the average sleep proportion between 1-2 am for the 96,645 UK Biobank participants that wore accelerometers on normal weekends.

We then reversed what Doherty et al. did so that we have 8 hours of sleep in a 23 hour day. (0.370833x 24 -0.9) / 23 = 0.347826.

0.347826 x 23 = 8 hours.

**Autumn**

The clocks go back an hour at 2am.

On the Saturday: You sleep from 10pm to midnight.

On the Sunday: You sleep from midnight to 6am (but it is now 5am). You then sleep from 10pm to midnight on the Sunday evening.

The hour from 1-2am has been recorded twice, but the accelerometer records you waking up at 5am instead of 6am so it still looks like you’ve slept for 8 hours:

12-1am, 1-2am, 1-2am, 2-3am, 3-4am, 4-5am, 10-11pm, 11-12pm

But this is out of a 25 hour day so sleep proportion = 8/25 = 0.32.

To turn it from a 25 hour day to a 24 hour day Doherty et al. removed the sleep for the hour 1-2am. e.g.(0.32 x 25-0.9) / 24 = 0.295833.

Doherty et al removed the data for the extra hour between 1-2am. Here we’ve calculated 0.9 as a comparative sleep proportion for the hour that Doherty subtracted. It is the average sleep proportion between 1-2 am for the 96,645 UK Biobank participants that wore accelerometers on normal weekends.

We then reversed what Doherty et al. did so that we have 8 hours sleep in a 25 hour day. (0.295833x 24+0.9) / 25 = 0.32.

0.32 x 25 = 8 hours.

**b) Assuming someone who normally sleeps for 8 hours (10pm-6am) but loses an hour’s sleep because they need to get up at their normal time on the Spring Sunday or gains an hour’s sleep because they take advantage of having a lie in on the Autumn Sunday.**

**Spring**

The clocks go forward an hour at 1am.

On the Saturday you sleep from 10pm to midnight.

On the Sunday you sleep from midnight to 5am (but it is now 6am). You then sleep from 10pm to midnight on the Sunday evening.

The hour from 1-2am does not exist in the accelerometer records, but it records you waking up at 6am instead of 5am so it records you sleeping for 7 hours on the Sunday:

12-1am, 2-3am, 3-4am, 4-5am, 5-6am, 10-11pm,11-12pm

But this is out of a 23 hour day so sleep proportion = 7/23 = 0.30434783

To turn it from a 23 hour day to a 24 hour day Doherty et al. imputed sleep for the hour 1-2am and added this in. e.g. (0.30434783 x 23 +0.9) / 24 = 0.32916667

Doherty et al used data from other days at the same time day of day to create the imputed value.

Here we’ve calculated 0.9 as a comparative sleep proportion for the hour that Doherty added. 0.9 is the average sleep proportion between 1-2 am for the 96,645 UK Biobank participants that wore accelerometers on normal weekends.

We then reversed what Doherty et al. did so that we have 7 hours of sleep in a 23 hour day. (0.32916667x 24 -0.9) / 23 = 0.30434783.

0.30434783 x 23 = 7 hours.

**Autumn**

The clocks go back an hour at 2am.

On the Saturday: You sleep from 10pm to midnight.

On the Sunday: You sleep from midnight to 7am (but it is now 6am). You then sleep from 10pm to midnight on the Sunday evening.

The hour from 1-2am has been recorded twice, but the accelerometer records you waking up at 6am instead of 7am so it still looks like you’ve slept for 9 hours:

12-1am, 1-2am, 1-2am, 2-3am, 3-4am, 4-5am, 5-6am, 10-11pm, 11-12pm

But this is out of a 25 hour day so sleep proportion = 9/25 = 0.36.

To turn it from a 25 hour day to a 24 hour day Doherty et al. removed the sleep for the hour 1-2am. e.g.(0.36 x 25-0.9) / 24 = 0.3375

Doherty et al removed the data for the extra hour between 1-2am. Here we’ve calculated 0.9 as a comparative sleep proportion for the hour that Doherty subtracted. It is the average sleep proportion between 1-2 am for the 96,645 UK Biobank participants that wore accelerometers on normal weekends.

We then reversed what Doherty et al. did so that we have 9 hours sleep in a 25 hour day. (0.3375 x 24+0.9) / 25 = 0.36.

0.36* 25 = 9 hours.
