## Supplementary material for "The effects of daylight saving time clock changes on accelerometer-measured sleep duration in the UK Biobank": Figures_S1-S10_SuppInfo

**Figures S1-S10** Mean daily sleep duration by day over the Spring and Autumn clock change fortnights, stratified by sociodemographics

**SPRING CLOCK CHANGE**

**Figure S1** Mean daily sleep duration by day over the Spring clock change by sex

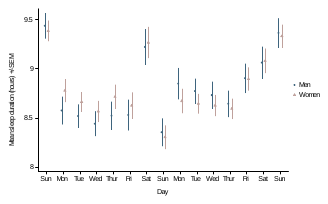

**Figure S2** Mean daily sleep duration by day over the Spring clock change by age

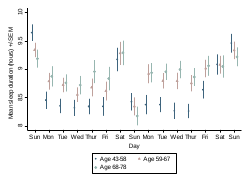

**Figure S3** Mean daily sleep duration by day over the Spring clock change by employment status

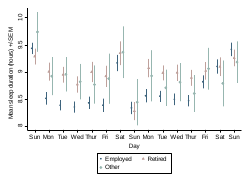

**Figure S4** Mean daily sleep duration by day over the Spring clock change by habitual sleep duration

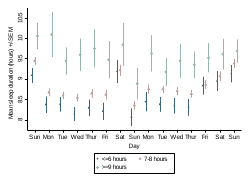

**Figure S5** Mean daily sleep duration by day over the Spring clock change by chronotype

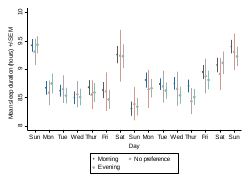

**AUTUMN CLOCK CHANGE**

**Figure S6** Mean daily sleep duration by day over the Autumn clock change by sex

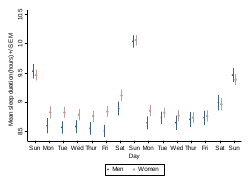

**Figure S7** Mean daily sleep duration by day over the Autumn clock change by age

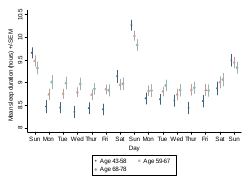

**Figure S8** Mean daily sleep duration by day over the Autumn clock change by employment status

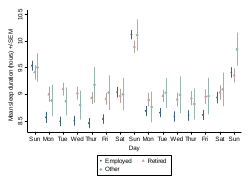

**Figure S9** Mean daily sleep duration by day over the Autumn clock change by habitual sleep duration

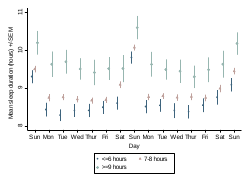

**Figure S10** Mean daily sleep duration by day over the Autumn clock change by chronotype

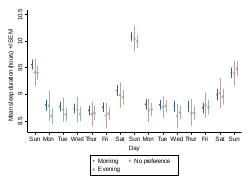
