## Supplementary material for "The effects of daylight saving time clock changes on accelerometer-measured sleep duration in the UK Biobank": Tables_S1-S4_SuppInfo

**Table S1** Number of participants by accelerometer start day and providing data for each day over the period of the clock changes

| **Start day** | **n spring** | **n autumn** | **Days with data** | | | | | | | | | | | | | | |
| --- | --- | --- | --- | --- | --- | --- | --- | --- | --- | --- | --- | --- | --- | --- | --- | --- | --- |
|  |  |  | **Sun**  **before** | **Mon**  **before** | **Tues before** | **Wed before** | **Thurs before** | **Fri before** | **Sat before** | **Sun clock change** | **Mon after** | **Tues**  **after** | **Wed**  **after** | **Thurs**  **after** | **Fri**  **after** | **Sat**  **after** | **Sun after** |
| 1.Mon 2 before | 403 | 432 | ✓ |  |  |  |  |  |  |  |  |  |  |  |  |  |  |
| 2. Wed 2 before | 243 | 422 | ✓ | ✓ | ✓ |  |  |  |  |  |  |  |  |  |  |  |  |
| 3. Thurs 2 before | 240 | 392 | ✓ | ✓ | ✓ | ✓ |  |  |  |  |  |  |  |  |  |  |  |
| 4. Fri 2 before | 493 | 441 | ✓ | ✓ | ✓ | ✓ | ✓ |  |  |  |  |  |  |  |  |  |  |
| 5. Sat 2 before | 406 | 590 | ✓ | ✓ | ✓ | ✓ | ✓ | ✓ |  |  |  |  |  |  |  |  |  |
| 6. Mon before | 379 | 694 |  |  | ✓ | ✓ | ✓ | ✓ | ✓ | ✓ |  |  |  |  |  |  |  |
| 7. Wed before | 103 | 433 |  |  |  |  | ✓ | ✓ | ✓ | ✓ | ✓ | ✓ |  |  |  |  |  |
| 8. Thurs before | 168 | 652 |  |  |  |  |  | ✓ | ✓ | ✓ | ✓ | ✓ | ✓ |  |  |  |  |
| 9. Friday before | 352 | 349 |  |  |  |  |  |  | ✓ | ✓ | ✓ | ✓ | ✓ | ✓ |  |  |  |
| 10. Sat before | 513 | 560 |  |  |  |  |  |  |  | ✓ | ✓ | ✓ | ✓ | ✓ | ✓ |  |  |
| 11. Mon after | 467 | 402 |  |  |  |  |  |  |  |  |  | ✓ | ✓ | ✓ | ✓ | ✓ | ✓ |
| 12. Wed after | 193 | 476 |  |  |  |  |  |  |  |  |  |  |  | ✓ | ✓ | ✓ | ✓ |
| 13. Thurs after | 262 | 502 |  |  |  |  |  |  |  |  |  |  |  |  | ✓ | ✓ | ✓ |
| 14. Fri after | 119 | 349 |  |  |  |  |  |  |  |  |  |  |  |  |  | ✓ | ✓ |
| 15. Sat after | 228 | 517 |  |  |  |  |  |  |  |  |  |  |  |  |  |  | ✓ |
|  |  | **n spring** | 1785 | 1382 | 1761 | 1518 | 1381 | 1056 | 1002 | 1515 | 1136 | 1603 | 1500 | 1525 | 1435 | 1041 | 1269 |
|  |  | **n autumn** | 2277 | 1845 | 2539 | 2117 | 2158 | 2369 | 2128 | 2688 | 1994 | 2396 | 1963 | 1787 | 1940 | 1729 | 2246 |

✓ represents days for which a person with a particular accelerometer start day has data. i.e. a person starting to wear the accelerometer on the second Monday before the clock change just has data for the Sunday before the clock change, whereas someone starting on the Monday before the clock change has data for the Tuesday before to the Sunday of the clock change.

**Table S2** Mean daily sleep duration (minutes) by day over the Spring & Autumn clock changes, overall and stratified by sociodemographics

|  | **Sunday before** | **Monday before** | **Tuesday before** | **Wednesday before** | **Thursday before** | **Friday before** | **Saturday before** | **Sunday of change** | **Monday after** | **Tuesday after** | **Wednesday after** | **Thursday after** | **Friday after** | **Saturday after** | **Sunday after** |
| --- | --- | --- | --- | --- | --- | --- | --- | --- | --- | --- | --- | --- | --- | --- | --- |
| **Spring (Mean (SE))** | | | | | | | | | | | | |  |  |  |
| Overall | 564.29 (2.48) | 521.28 (2.76) | 515.84 (2.33) | 510.56 (2.48) | 517.68 (2.85) | 514.77 (3.10) | 554.63 (3.63) | 499.42 (2.79) | 524.73 (2.99) | 521.77 (2.41) | 520.09 (2.59) | 516.74 (2.49) | 533.78  (2.82) | 544.17 (3.06) | 560.56 (2.83) |
| Sex | | | | | | | | | | | | | | | |
| Female | 562.97 (3.18) | 526.53 (3.57) | 519.62 (3.05) | 513.86 (3.24) | 522.80 (3.76) | 517.42 (4.09) | 555.88 (4.79) | 498.12 (3.66) | 520.20 (3.77) | 518.56 (3.05) | 517.45 (3.25) | 515.47 (3.11) | 533.62 (3.58) | 544.66 (3.80) | 559.74 (3.58) |
| Male | 566.00 (3.94) | 514.41 (4.31) | 510.98 (3.60) | 506.35 (3.84) | 511.16 (4.36) | 511.60 (4.74) | 553.11 (5.55) | 501.10 (4.29) | 530.76 (4.83) | 526.15 (3.87) | 523.70 (4.22) | 518.47 (4.08) | 534.01 (4.53) | 543.54 (5.00) | 561.65 (4.58) |
| Age | | | | | | | | | | | | |  |  |  |
| Tertile 1 (age 43-59) | 578.45 (4.37) | 507.22 (4.58) | 501.17 (3.77) | 499.12 (4.24) | 500.54 (4.11) | 500.63 (4.93) | 550.14 (6.28) | 505.54 (4.67) | 502.49 (5.11) | 502.54 (4.02) | 496.08 (4.30) | 495.74 (3.79) | 518.19 (4.68) | 544.89 (5.42) | 567.33 (5.02) |
| Tertile 2 (age 60-67) | 560.20 (3.80) | 527.08 (4.43) | 523.30 (3.84) | 513.08 (3.84) | 520.62 (4.76) | 516.86 (5.02) | 556.66 (6.07) | 500.67 (4.76) | 535.09 (5.03) | 527.67 (4.04) | 527.51 (4.31) | 525.10 (4.31) | 540.69 (4.65) | 544.62 (4.75) | 559.85 (4.64) |
| Tertile 3 (age 68-77) | 550.95 (4.76) | 532.37 (5.39) | 525.49 (4.56) | 523.15 (4.89) | 537.34 (6.12) | 530.03 (6.26) | 557.24 (6.53) | 490.58 (5.00) | 536.15 (5.17) | 537.23 (4.31) | 539.58 (4.62) | 531.66 (4.70) | 543.47 (5.31) | 542.77 (5.87) | 552.89 (5.01) |
| Chronotype (self-reported) | | | | | | | | | | | | | | | |
| Morning | 564.98 (3.29) | 520.25 (3.62) | 517.19 (3.04) | 509.80 (3.31) | 520.67 (3.93) | 517.87 (4.26) | 555.27 (4.56) | 498.35 (3.82) | 528.50 (4.02) | 524.13 (3.07) | 524.98 (3.36) | 522.25 (3.40) | 536.96 (3.69) | 544.30 (3.85) | 563.75 (3.57) |
| No preference | 558.26 (7.01) | 515.05 (6.58) | 518.92 (7.33) | 513.51 (7.05) | 513.09 (7.06) | 516.36 (10.30) | 554.19 (13.90) | 503.45 (8.91) | 519.11 (10.10) | 521.57 (8.57) | 519.26 (9.06) | 506.34 (6.86) | 532.80 (9.31) | 534.90 (8.58) | 558.32 (9.76) |
| Evening | 565.66 (4.53) | 524.77 (5.28) | 512.04 (4.17) | 510.46 (4.44) | 515.38 (4.97) | 508.00 (5.04) | 553.61 (6.38) | 500.36 (4.64) | 519.87 (4.96) | 517.40 (4.30) | 512.46 (4.54) | 510.67 (4.31) | 528.59 (5.01) | 546.62 (6.00) | 553.36 (5.22) |
| Sleep duration (self-reported) | | | | | | | | | | | | | | | |
| <=6 hours | 545.20 (5.42) | 502.34 (6.02) | 502.68 (5.34) | 488.41 (5.12) | 497.49 (6.13) | 492.56 (6.38) | 551.03 (8.52) | 483.72 (6.57) | 506.62 (7.18) | 502.56 (5.41) | 500.69 (5.76) | 498.15 (6.13) | 530.01 (6.58) | 536.69 (7.43) | 547.46 (6.41) |
| 7-8 hours | 566.47 (2.87) | 520.22 (2.89) | 515.72 (2.62) | 512.45 (2.81) | 518.76 (3.22) | 516.95 (3.62) | 552.79 (4.13) | 501.37 (3.17) | 525.17 (3.31) | 524.84 (2.75) | 522.08 (2.91) | 518.06 (2.77) | 531.73 (3.23) | 543.91 (3.44) | 562.80 (3.32) |
| >=9 hours | 603.17 (9.77) | 605.13 (16.80) | 566.39 (10.14) | 575.59 (12.11) | 584.76 (14.19) | 568.23 (13.85) | 590.40 (16.06) | 533.09 (11.73) | 577.48 (13.46) | 550.29 (10.56) | 566.55 (12.58) | 561.03 (9.81) | 571.09 (10.91) | 576.47 (11.44) | 580.93 (8.69) |
| Current employment status | | | | | | | | | | | | | | | |
| Employed | 565.93 (3.12) | 510.61 (3.43) | 502.99 (2.80) | 500.99 (3.09) | 505.72 (3.37) | 503.02 (3.69) | 549.75 (4.52) | 499.99 (3.42) | 513.47 (3.71) | 512.91 (3.05) | 509.53 (3.24) | 508.00 (3.11) | 528.97 (3.60) | 545.86 (3.95) | 564.82 (3.74) |
| Retired | 556.88 (4.39) | 540.17 (5.02) | 536.74 (4.35) | 525.98 (4.49) | 539.81 (5.64) | 535.05 (5.90) | 560.63 (6.64) | 496.55 (5.03) | 544.06 (5.09) | 538.74 (4.15) | 539.07 (4.53) | 533.33 (4.41) | 541.05 (4.90) | 545.66 (5.18) | 555.40 (4.67) |
| Other | 584.17 (11.13) | 535.25 (10.74) | 537.29 (9.68) | 529.03 (9.98) | 525.89 (10.73) | 532.40 (14.07) | 561.79 (14.50) | 506.33 (13.02) | 535.53 (14.33) | 522.34 (10.18) | 528.56 (10.70) | 515.96 (10.27) | 543.46 (11.69) | 527.17 (12.90) | 550.76 (11.68) |
| **Autumn (Mean (SE))** | | | | | | | | | | | | | | | |
| Overall | 569.42 (2.23) | 523.72 (2.44) | 522.90 (2.11) | 521.80 (2.16) | 520.20 (2.16) | 521.76 (2.06) | 541.07 (2.31) | 602.82 (2.06) | 525.89 (2.19) | 527.02 (2.00) | 523.16 (2.29) | 523.46 (2.31) | 525.14 (2.31) | 538.79 (2.49) | 565.31 (2.25) |
| Sex | | | | | | | | | | | | | | | |
| Female | 567.72 (2.75) | 529.72 (3.12) | 529.76 (2.81) | 527.20 (2.84) | 525.61 (2.85) | 530.68 (2.77) | 547.19 (3.00) | 603.33 (2.71) | 531.32 (2.88) | 529.38 (2.56) | 526.35 (2.89) | 524.23 (2.86) | 525.98 (2.92) | 538.16 (3.13) | 563.32 (2.88) |
| Male | 571.64 (3.67) | 515.93 (3.87) | 514.14 (3.16) | 514.98 (3.30) | 513.37 (3.29) | 510.65 (3.05) | 533.58 (3.57) | 602.17 (3.16) | 518.91 (3.36) | 523.98 (3.17) | 519.01 (3.69) | 522.49 (3.79) | 524.08 (3.70) | 539.54 (3.99) | 567.72 (3.53) |
| Age | | | | | | | | | | | | | | | |
| Tertile 1 (age 43-59) | 579.1 (3.61) | 508.17 (4.29) | 506.82 (3.63) | 501.14 (3.91) | 506.04 (3.82) | 504.56 (3.75) | 548.00 (4.19) | 615.54 (3.61) | 518.95 (3.79) | 517.80 (3.47) | 515.92 (3.94) | 506.48 (4.04) | 515.47 (4.04) | 532.21 (4.55) | 569.23 (4.11) |
| Tertile 2 (age 60-67) | 568.31 (3.72) | 524.34 (3.80) | 525.11 (3.20) | 527.33 (3.21) | 523.82 (3.50) | 530.98 (3.07) | 537.13 (3.67) | 601.63 (3.29) | 529.19 (3.51) | 528.04 (3.17) | 524.26 (3.82) | 530.48 (3.90) | 530.18 (3.90) | 540.26 (3.97) | 566.46 (3.61) |
| Tertile 3 (age 68-77) | 559.10 (4.28) | 540.54 (4.63) | 539.20 (4.19) | 538.19 (4.10) | 531.53 (3.85) | 529.64 (3.94) | 538.41 (4.18) | 589.38 (3.81) | 529.39 (4.14) | 536.04 (3.80) | 529.86 (4.15) | 533.04 (3.98) | 529.43 (4.02) | 543.75 (4.48) | 559.49 (3.98) |
| Chronotype (self-reported) | | | | | | | | | | | | | | | |
| Morning | 573.23 (2.86) | 527.53 (3.10) | 525.96 (2.75) | 523.68 (2.73) | 521.49 (2.74) | 525.11 (2.73) | 543.98 (3.02) | 604.85 (2.64) | 528.44 (2.97) | 527.96 (2.64) | 526.52 (3.11) | 526.40 (3.11) | 524.48 (3.03) | 539.28 (3.20) | 564.25 (2.88) |
| No preference | 564.44 (7.95) | 526.92 (8.68) | 522.60 (6.90) | 522.54 (7.45) | 517.38 (8.68) | 515.73 (7.10) | 538.94 (7.19) | 602.55 (7.31) | 521.59 (6.61) | 525.41 (6.09) | 515.59 (6.88) | 519.17 (7.69) | 527.63 (7.14) | 541.22 (8.62) | 563.62 (7.21) |
| Evening | 564.03 (4.02) | 515.36 (4.48) | 517.16 (3.79) | 517.66 (4.00) | 518.67 (4.08) | 517.77 (3.54) | 536.54 (4.19) | 599.36 (3.69) | 522.82 (3.75) | 526.56 (3.60) | 519.27 (3.93) | 518.61 (3.86) | 525.25 (4.17) | 537.10 (4.50) | 568.58 (4.22) |
| Sleep duration (self-reported) | | | | | | | | | | | | | | | |
| <=6 hours | 612.03 (5.15) | 505.83 (5.47) | 497.07 (4.63) | 504.57 (4.99) | 504.62 (5.17) | 509.42 (5.07) | 516.24 (5.13) | 588.28 (4.77) | 510.63 (5.24) | 512.64 (5.03) | 504.41 (5.86) | 502.60 (5.31) | 512.85 (5.53) | 525.32 (5.64) | 599.26 (5.28) |
| 7-8 hours | 623.85 (2.52) | 524.83 (2.78) | 525.58 (2.37) | 522.31 (2.44) | 520.29 (2.38) | 521.24 (2.28) | 545.37 (2.60) | 604.09 (2.33) | 526.09 (2.41) | 527.44 (2.22) | 524.97 (2.51) | 526.31 (2.65) | 524.57 (2.59) | 539.24 (2.86) | 620.75 (2.55) |
| >=9 hours | 665.64 (9.40) | 577.71 (10.20) | 581.64 (9.46) | 570.26 (8.59) | 564.70 (10.14) | 570.80 (9.06) | 570.91 (10.79) | 635.57 (9.30) | 577.63 (9.90) | 569.35 (8.13) | 566.68 (9.29) | 557.84 (8.99) | 569.03 (9.93) | 577.95 (10.72) | 664.56 (8.94) |
| Current employment status | | | | | | | | | | | | | | | |
| Employed | 572.05 (2.81) | 513.92 (3.09) | 509.36 (2.67) | 510.30 (2.75) | 507.51 (2.80) | 512.01 (2.65) | 542.30 (3.00) | 606.97 (2.63) | 521.17 (2.79) | 519.33 (2.55) | 514.89 (2.87) | 516.12 (2.98) | 516.72 (2.85) | 536.25 (3.25) | 564.55 (2.86) |
| Retired | 564.61 (4.11) | 539.84 (4.37) | 545.79 (3.64) | 541.22 (3.68) | 535.84 (3.37) | 534.83 (3.29) | 538.19 (3.88) | 593.36 (3.48) | 533.90 (3.81) | 538.13 (3.42) | 534.13 (3.92) | 535.47 (4.00) | 537.36 (4.08) | 541.86 (4.24) | 561.17 (3.89) |
| Other | 570.21 (8.09) | 532.90 (8.96) | 532.07 (7.93) | 527.80 (8.28) | 550.63 (10.19) | 541.78 (9.85) | 539.90 (9.21) | 606.56 (8.97) | 525.65 (8.80) | 541.66 (8.42) | 539.31 (10.49) | 529.10 (8.74) | 537.94 (10.43) | 545.59 (9.55) | 590.67 (9.46) |

**Table S3** Between-individual comparisons of mean daily sleep duration (minutes) on the Sunday of the clock change versus the Sunday before and the Sunday after.

|  | **Sun clock change** | | Sunday before | | | |  |  | | **Sunday after** | | | |
| --- | --- | --- | --- | --- | --- | --- | --- | --- | --- | --- | --- | --- | --- |
|  | **n** | **Mean sleep duration in minutes (SD)** | **n** | **Mean sleep duration in minutes (SD)** | **Difference: Sun of clock change to Sun before (SE)** | **p value** | **Cochran’s Q p value** | **n** | **Mean sleep duration in minutes (SD)** | | **Difference: Sun of clock change to Sun after (SE)** | **p value** | **Cochran’s Q** |
| **Spring** | | | | | | | | | | | | | |
| Overall | 1515 | 499.42 (108.42) | 1785 | 564.29 (104.91) | -64.87 (3.72) | 3.667x10^-65^ |  | 1269 | 560.56 (100.96) | | -61.14 (4.00) | 9.902x10^-51^ |  |
| Sex | | | | | | | | | | | | | |
| Female | 857 | 498.12 (107.28) | 1010 | 562.97 (101.02) | -64.85 (4.83) | 2.447x10^-39^ | 0.994 | 727 | 559.74 (96.59) | | -61.61 (5.17) | 1.971x10^-31^ | 0.896 |
| Male | 658 | 501.10 (109.95) | 775 | 566.00 (109.81) | -64.90 (5.82) | 1.030x10-^27^ |  | 542 | 561.65 (106.63) | | -60.55 (6.29) | 3.590x10^-21^ |  |
| Age | | | | | | | | | | | | | |
| Tertile 1  (age 43-59) | 511 | 505.54 (105.54) | 641 | 578.45 (110.64) | -72.91 (6.43) | 2.487x10^-28^ | 0.254 | 447 | 567.33 (106.21) | | -61.79 (6.86) | 1.062x10^-18^ | 0.942 |
| Tertile 2  (age 60-67) | 569 | 500.67 (113.66) | 668 | 560.20 (98.29) | -59.53 (6.03) | 3.336x10^-22^ |  | 470 | 559.85 (100.61) | | -59.19 (6.73) | 5.808x10^-18^ |  |
| Tertile 3  (age 68-77) | 435 | 490.58 (104.34) | 476 | 550.95 (103.95) | -60.37 (6.91) | 1.113x10^-17^ |  | 352 | 552.89 (94.07) | | -62.30 (7.16) | 1.916x10^-17^ |  |
| Chronotype (self-reported) | | | | | | | | | | | | | |
| Morning | 820 | 498.35 (109.28) | 994 | 564.98 (103.65) | -66.63 (5.01) | 1.474x10^-38^ | 0.626 | 744 | 563.75 (97.37) | | -65.40 (5.25) | 5.651x10^-34^ | 0.334 |
| No preference | 156 | 503.45 (111.27) | 194 | 558.26 (97.66) | -54.81 (11.18) | 1.445x10^-06^ |  | 123 | 558.32 (108.29) | | -54.88 (13.26) | 4.644x10^-05^ |  |
| Evening | 529 | 500.36 (106.72) | 585 | 565.66 (109.64) | -65.30 (6.50) | 8.099x10^-23^ |  | 393 | 553.36 (103.51) | | -53.00 (7.02) | 1.026x10^-13^ |  |
| Sleep duration (self-reported) | | | | | | | | | | | | | |
| <=6 hours | 316 | 483.72 (116.80 | 403 | 545.20 (108.79) | -61.48 (8.44) | 8.771x10^-13^ | 0.870 | 280 | 547.46 (107.23) | | -63.74 (9.23) | 1.265x10^-11^ | 0.651 |
| 7-8 hours | 1104 | 501.37 (105.37 | 1252 | 566.47 (101.70) | -65.10 (4.27) | 4.092x10^-50^ |  | 914 | 562.80 (100.38) | | -61.43 (4.61) | 7.490x10^-39^ |  |
| >=9 hours | 87 | 533.09 (109.40) | 124 | 603.17 (108.82) | -70.08 (15.25) | 7.477x10^-06^ |  | 73 | 580.93 (74.21) | | -47.84 (15.08) | 1.813x10^-03^ |  |
| Current employment status (self-reported) | | | | | | | | | | | | | |
| Employed | 931 | 499.99 (104.43) | 1133 | 565.93 (104.94) | -65.94 (4.63) | 6.041x10^-44^ | 0.579 | 751 | 564.82 (102.36) | | -64.83 (5.08) | 1.046x10^-35^ | 0.478 |
| Retired | 464 | 496.55 (108.29) | 537 | 556.88 (101.81) | -60.33 (6.65) | 5.835x10^-19^ |  | 425 | 555.40 (96.24) | | -58.85 (6.90) | 6.088x10^-17^ |  |
| Other | 113 | 506.33 (138.37) | 111 | 584.17 (117.27) | -77.85 (17.15) | 9.275x10^-06^ |  | 89 | 550.76 (110.19) | | -44.43 (17.96) | 1.421x10^-02^ |  |
| **Autumn** | | | | | | | | | | | | | |
| Overall | 2688 | 602.82 (106.75) | 2277 | 569.42 (106.42) | 33.39 (3.04) | 8.065x10^-28^ |  | 2246 | 565.31 (106.45) | | 37.51 (3.05) | 2.677x10^-34^ |  |
| Sex | | | | | | | | | | | | | |
| Female | 1503 | 603.33 (105.09) | 1289 | 567.72 (98.91) | 35.60 (3.88) | 8.968x10^-20^ | 0.412 | 1229 | 563.32 (101.06) | | 40.00 (3.97) | 1.915x10^-23^ | 0.369 |
| Male | 1185 | 602.17 (108.86) | 988 | 571.64 (115.50) | 30.53 (4.82) | 2.946x10^-10^ |  | 1017 | 567.72 (112.61) | | 34.46 (4.73) | 4.383x10^-13^ |  |
| Age | | | | | | | | | | | | | |
| Tertile 1  (age 43-59) | 899 | 615.54 (108.15) | 771 | 579.10 (100.36) | 36.44 (5.14) | 1.907x10^-12^ | 0.724 | 719 | 569.23 (110.26) | | 46.31 (5.46) | 4.852x10^-17^ | 0.095 |
| Tertile 2  (age 60-67) | 1029 | 601.63 (105.57) | 878 | 568.31 (110.35) | 33.33 (4.95) | 2.251x10^-11^ |  | 872 | 566.46 (106.49) | | 35.18 (4.88) | 8.008x10^-13^ |  |
| Tertile 3  (age 68-77) | 760 | 589.38 (105.06) | 628 | 559.10 (107.17) | 30.28 (5.72) | 1.379x10^-07^ |  | 655 | 559.49 (101.94) | | 29.89 (5.52) | 7.405x10^-08^ |  |
| Chronotype (self-reported) | | | | | | | | | | | | | |
| Morning | 1519 | 604.85 (102.79) | 1322 | 573.23 (103.93) | 31.62 (3.89) | 6.079x10^-16^ | 0.770 | 1308 | 564.25 (104.09) | | 40.60 (3.90) | 6.320x10^-25^ | 0.352 |
| No preference | 278 | 602.55 (121.86) | 223 | 564.44 (118.71) | 38.11 (10.83) | 4.734x10^-04^ |  | 235 | 563.62 (110.46) | | 38.92 (10.35) | 1.884x10^-04^ |  |
| Evening | 871 | 599.36 (109.01) | 716 | 564.03 (107.50) | 35.33 (5.46) | 1.343x10^-10^ |  | 683 | 568.58 (110.29) | | 30.79 (5.60) | 4.501x10^-08^ |  |
| Sleep duration (self-reported) | | | | | | | | | | | | | |
| <=6 hours | 535 | 588.28 (110.22) | 525 | 558.03 (117.95) | 30.25 (7.01) | 1.749x10^-05^ | 0.688 | 460 | 545.26 (113.33) | | 43.02 (7.10) | 1.941x10^-09^ | 0.464 |
| 7-8 hours | 1973 | 604.09 (103.66) | 1602 | 569.85 (100.83) | 34.24 (3.44) | 5.422x10^-23^ |  | 1622 | 566.75 (102.50) | | 37.34 (3.46) | 8.623x10^-27^ |  |
| >=9 hours | 168 | 635.57 (120.51) | 141 | 611.64 (111.67) | 23.93 (13.31) | 7.325x10^-02^ |  | 155 | 610.56 (111.36) | | 25.01 (12.94) | 5.418x10^-02^ |  |
| Current employment status (self-reported) | | | | | | | | | | | | | |
| Employed | 1607 | 606.97 (105.46) | 1376 | 572.05 (104.27) | 34.92 (3.85) | 2.280x10^-19^ | 0.621 | 1343 | 564.55 (104.78) | | 42.42 (3.89) | 3.308x10^-27^ | 0.069 |
| Retired | 878 | 593.36 (103.19) | 723 | 564.61 (110.44) | 28.75 (5.35) | 8.808x10^-08^ |  | 742 | 561.17 (106.08) | | 32.19 (5.21) | 8.334x10^-10^ |  |
| Other | 196 | 606.56 (125.58) | 174 | 570.21 (106.67) | 36.36 (12.19) | 3.060x10^-03^ |  | 159 | 590.67 (119.26) | | 15.89 (13.11) | 2.261x10^-01^ |  |

**Table S4** Between-individual comparisons of mean sleep duration (hours and minutes) on the Monday to Saturday before and after the Spring and Autumn clock changes, overall and stratified by sociodemographics

| **Spring** | | | |  | **Autumn** | | |  |
| --- | --- | --- | --- | --- | --- | --- | --- | --- |
|  | **Difference in sleep duration (minutes)** | **95% CI** | **p value** | **Cochran’s Q p value*** | **Difference in sleep duration**  **(minutes)** | **95% CI** | **p value** | **Cochran’s Q p value*** |
| **Overall** | | | |  | **Overall** | | |  |
| Mon after vs before | 3.45 | -4.52,11.42 | 0.396 |  | 2.17 | -4.26,8.61 | 0.508 |  |
| Tue after vs before | 5.93 | -0.65,12.50 | 0.077 |  | 4.12 | -1.57,9.81 | 0.156 |  |
| Wed after vs before | 9.53 | 2.51,16.56 | 0.008 |  | 1.36 | -4.81,7.53 | 0.666 |  |
| Thur after vs before | -0.95 | -8.37,6.48 | 0.803 |  | 3.26 | -2.94,9.46 | 0.303 |  |
| Fri after vs before | 19.01 | 10.80,27.22 | 5.945x10^-06^ |  | 3.38 | -2.68,9.45 | 0.274 |  |
| Sat after vs before | -10.46 | -19.76,-1.16 | 0.028 |  | -2.28 | -8.94,4.38 | 0.502 |  |
| **Sex** | | | | | **Sex** | | | |
| *Female* | | | |  | *Female* | | |  |
| Mon after vs before | -6.33 | -16.51,3.85 | 0.223 | 0.006 | 1.60 | -6.72,9.93 | 0.706 | 0.835 |
| Tues after vs before | -1.07 | -9.54,7.40 | 0.805 | 0.017 | -0.38 | -7.83,7.07 | 0.920 | 0.082 |
| Wed after vs before | 3.59 | -5.40,12.58 | 0.434 | 0.060 | -0.85 | -8.79,7.09 | 0.834 | 0.446 |
| Thurs after vs before | -7.33 | -16.89,2.24 | 0.133 | 0.058 | -1.39 | -9.31,6.54 | 0.732 | 0.103 |
| Fri after vs before | 16.19 | 5.54,26.85 | 0.003 | 0.466 | -4.70 | -12.60,3.19 | 0.243 | 0.004 |
| Sat after vs before | -11.22 | -23.22,0.78 | 0.067 | 0.864 | -9.02 | -17.53,-0.52 | 0.038 | 0.030 |
| *Male* | | | |  | *Male* | | |  |
| Mon after vs before | 16.35 | 3.66,29.05 | 0.012 |  | 2.99 | -7.07,13.04 | 0.560 |  |
| Tues after vs before | 15.18 | 4.80,25.55 | 0.004 |  | 9.84 | 1.05,18.62 | 0.028 |  |
| Wed after vs before | 17.35 | 6.15,28.55 | 0.002 |  | 4.03 | -5.69,13.74 | 0.416 |  |
| Thurs after vs before | 7.30 | -4.41,19.02 | 0.222 |  | 9.11 | -0.73,18.95 | 0.070 |  |
| Fri after vs before | 22.41 | 9.54,35.28 | 0.001 |  | 13.43 | 4.03,22.82 | 0.005 |  |
| Sat after vs before | -9.57 | -24.23,5.08 | 0.200 |  | 5.96 | -4.54,16.46 | 0.266 |  |
| **Age** | | | | | **Age** | | | |
| *Tertile 1 (age 43-58)* | | | |  | *Tertile 1 (age 43-58)* | | |  |
| Mon after vs before | -4.73 | -18.19,8.73 | 0.491 | 0.404 | 10.78 | -0.45,22.01 | 0.060 | 0.029 |
| Tues after vs before | 1.37 | -9.45,12.19 | 0.804 | 0.453 | 10.98 | 1.13,20.83 | 0.029 | 0.167 |
| Wed after vs before | -3.04 | -14.89,8.80 | 0.614 | 0.048 | 14.78 | 3.89,25.67 | 0.008 | 0.009 |
| Thurs after vs before | -4.80 | -15.78,6.17 | 0.391 | 0.474 | 0.44 | -10.47,11.34 | 0.938 | 0.679 |
| Fri after vs before | 17.57 | 4.22,30.91 | 0.010 | 0.608 | 10.91 | 0.10,21.72 | 0.048 | 0.227 |
| Sat after vs before | -5.25 | -21.52,11.02 | 0.527 | 0.725 | -15.79 | -27.92,-3.66 | 0.011 | 0.026 |
| *Tertile 2 (age 59-67)* | | | |  | *Tertile 2 (age 59-67)* | | |  |
| Mon after vs before | 8.02 | -5.13,21.16 | 0.232 |  | 4.85 | -5.30,14.99 | 0.349 |  |
| Tues after vs before | 4.36 | -6.58,15.31 | 0.435 |  | 2.94 | -5.90,11.77 | 0.515 |  |
| Wed after vs before | 14.43 | 3.10,25.76 | 0.013 |  | -3.07 | -12.85,6.70 | 0.538 |  |
| Thurs after vs before | 4.48 | -8.12,17.09 | 0.486 |  | 6.67 | -3.61,16.95 | 0.204 |  |
| Fri after vs before | 23.82 | 10.41,37.23 | 0.001 |  | -0.80 | -10.54,8.94 | 0.872 |  |
| Sat after vs before | -12.04 | -27.16,3.08 | 0.119 |  | 3.13 | -7.46,13.73 | 0.562 |  |
| *Tertile 3 (age 68-78)* | | | |  | *Tertile 3 (age 68-78)* | | |  |
| Mon after vs before | 3.78 | -10.87,18.43 | 0.612 |  | -11.15 | -23.33,1.04 | 0.073 |  |
| Tues after vs before | 11.73 | -0.58,24.05 | 0.062 |  | -3.15 | -14.24,7.94 | 0.577 |  |
| Wed after vs before | 16.43 | 3.22,29.63 | 0.015 |  | -8.33 | -19.77,3.12 | 0.154 |  |
| Thurs after vs before | -5.69 | -20.84,9.47 | 0.462 |  | 1.51 | -9.36,12.37 | 0.786 |  |
| Fri after vs before | 13.44 | -2.66,29.54 | 0.102 |  | -0.21 | -11.25,10.83 | 0.970 |  |
| Sat after vs before | -14.47 | -31.70,2.75 | 0.100 |  | 5.34 | -6.68,17.36 | 0.384 |  |
| **Chronotype (self-reported)** | | | |  | **Chronotype (self-reported)** | | |  |
| *Morning* | | | |  | *Morning* | | |  |
| Mon after vs before | 8.25 | -2.36,18.86 | 0.128 | 0.349 | 0.91 | -7.51,9.34 | 0.832 | 0.505 |
| Tues after vs before | 6.95 | -1.53,15.42 | 0.108 | 0.929 | 2.00 | -5.47,9.47 | 0.600 | 0.510 |
| Wed after vs before | 15.17 | 5.92,24.43 | 0.001 | 0.232 | 2.84 | -5.27,10.95 | 0.492 | 0.672 |
| Thurs after vs before | 1.58 | -8.61,11.78 | 0.7601 | 0.655 | 4.91 | -3.22,13.05 | 0.236 | 0.770 |
| Fri after vs before | 19.09 | 8.03,30.15 | 0.001 | 0.962 | -0.63 | -8.64,7.38 | 0.878 | 0.328 |
| Sat after vs before | -10.97 | -22.68,0.74 | 0.066 | 0.796 | -4.70 | -13.34,3.93 | 0.286 | 0.713 |
| *No preference* | | | |  | *No preference* | | |  |
| Mon after vs before | 4.06 | -19.64,27.77 | 0.736 |  | -5.33 | -26.76,16.10 | 0.625 |  |
| Tues after vs before | 2.65 | -19.55,24.85 | 0.815 |  | 2.81 | -15.29,20.90 | 0.761 |  |
| Wed after vs before | 5.75 | -16.85,28.35 | 0.617 |  | -6.95 | -26.88,12.97 | 0.493 |  |
| Thurs after vs before | -6.75 | -26.99,13.48 | 0.512 |  | 1.79 | -18.45,22.04 | 0.862 |  |
| Fri after vs before | 16.44 | -10.86,43.74 | 0.237 |  | 11.90 | -7.89,31.69 | 0.238 |  |
| Sat after vs before | -19.30 | -51.39,12.80 | 0.238 |  | 2.28 | -19.77,24.33 | 0.839 |  |
| *Evening* | | | |  | *Evening* | | |  |
| Mon after vs before | -4.89 | -19.12,9.34 | 0.500 |  | 7.45 | -4.02,18.92 | 0.203 |  |
| Tues after vs before | 5.36 | -6.39,17.10 | 0.371 |  | 9.41 | -0.84,19.65 | 0.072 |  |
| Wed after vs before | 2.00 | -10.46,14.45 | 0.753 |  | 1.60 | -9.40,12.61 | 0.775 |  |
| Thurs after vs before | -4.71 | -17.62,8.21 | 0.475 |  | -0.06 | -11.08,10.95 | 0.991 |  |
| Fri after vs before | 20.60 | 6.64,34.55 | 0.004 |  | 7.48 | -3.25,18.22 | 0.172 |  |
| Sat after vs before | -6.99 | -24.17,10.19 | 0.425 |  | 0.56 | -11.50,12.62 | 0.927 |  |
| **Sleep duration (self-reported)** | | | |  | **Sleep duration (self-reported)** | | |  |
| *<=6 hours* | | | |  | *<=6 hours* | | |  |
| Mon after vs before | 4.28 | -14.12,22.67 | 0.648 | 0.336 | 4.80 | -10.06,19.66 | 0.526 | 0.907 |
| Tues after vs before | -0.12 | -15.06,14.82 | 0.987 | 0.169 | 15.57 | 2.15,28.99 | 0.023 | 0.084 |
| Wed after vs before | 12.28 | -2.85,27.41 | 0.111 | 0.538 | -0.15 | -15.26,14.95 | 0.984 | 0.859 |
| Thurs after vs before | 0.66 | -16.37,17.68 | 0.940 | 0.420 | -2.03 | -16.58,12.52 | 0.784 | 0.445 |
| Fri after vs before | 37.45 | 19.45,55.45 | 5.05x10^-05^ | 0.060 | 3.43 | -11.29,18.15 | 0.648 | 0.934 |
| Sat after vs before | -14.34 | -36.52,7.85 | 0.205 | 0.891 | 9.08 | -5.88,24.03 | 0.234 | 0.167 |
| *7-8 hours* | | | |  | *7-8 hours* | | |  |
| Mon after vs before | 4.95 | -3.67,13.58 | 0.260 |  | 1.26 | -5.96,8.47 | 0.733 |  |
| Tues after vs before | 9.12 | 1.67,16.58 | 0.017 |  | 1.86 | -4.52,8.23 | 0.568 |  |
| Wed after vs before | 9.63 | 1.70,17.56 | 0.017 |  | 2.66 | -4.20,9.52 | 0.448 |  |
| Thurs after vs before | -0.70 | -9.03,7.64 | 0.870 |  | 6.02 | -0.97,13.01 | 0.091 |  |
| Fri after vs before | 14.78 | 5.27,24.28 | 0.002 |  | 3.33 | -3.44,10.10 | 0.335 |  |
| Sat after vs before | -8.87 | -19.41,1.66 | 0.099 |  | -6.13 | -13.71,1.45 | 0.113 |  |
| *>=9 hours* | | | | | *>=9 hours* | | | |
| Mon after vs before | -27.65 | -70.11,14.81 | 0.201 |  | -0.08 | -28.03,27.87 | 0.996 |  |
| Tues after vs before | -16.10 | -45.03,12.83 | 0.274 |  | -12.29 | -36.85,12.27 | 0.326 |  |
| Wed after vs before | -9.04 | -43.53,25.45 | 0.606 |  | -3.58 | -28.47,21.30 | 0.777 |  |
| Thurs after vs before | -23.73 | -57.77,10.32 | 0.171 |  | -6.86 | -33.52,19.80 | 0.613 |  |
| Fri after vs before | 2.86 | -31.88,37.60 | 0.871 |  | -1.77 | -28.22,24.68 | 0.896 |  |
| Sat after vs before | -13.93 | -52.74,24.88 | 0.480 |  | 7.03 | -22.88,36.95 | 0.644 |  |
| **Current employment status** | | | |  | **Current employment status** | | |  |
| *Employed* | | | | | *Employed* | | | |
| Mon after vs before | 2.86 | -7.04,12.76 | 0.571 | 0.981 | 7.25 | -0.91,15.42 | 0.082 | 0.135 |
| Tues after vs before | 9.92 | 1.80,18.04 | 0.017 | 0.171 | 9.98 | 2.73,17.22 | 0.007 | 0.016 |
| Wed after vs before | 8.54 | -0.24,17.32 | 0.057 | 0.664 | 4.59 | -3.21,12.39 | 0.249 | 0.158 |
| Thurs after vs before | 2.29 | -6.71,11.28 | 0.618 | 0.484 | 8.61 | 0.59,16.64 | 0.036 | 0.062 |
| Fri after vs before | 25.95 | 15.84,36.06 | 5.437e-07 | 0.088 | 4.71 | -2.92,12.34 | 0.226 | 0.823 |
| Sat after vs before | -3.89 | -15.66,7.88 | 0.517 | 0.229 | -6.05 | -14.72,2.62 | 0.172 | 0.342 |
| *Retired* | | | | | *Retired* | | | |
| Mon after vs before | 3.89 | -10.14,17.93 | 0.586 |  | -5.95 | -17.32,5.42 | 0.305 |  |
| Tues after vs before | 2.00 | -9.81,13.80 | 0.740 |  | -7.67 | -17.46,2.13 | 0.125 |  |
| coWed after vs before | 13.09 | 0.57,25.61 | 0.040 |  | -7.08 | -17.62,3.45 | 0.188 |  |
| Thurs after vs before | -6.48 | -20.54,7.58 | 0.366 |  | -0.37 | -10.61,9.88 | 0.944 |  |
| Fri after vs before | 6.00 | -9.05,21.05 | 0.434 |  | 2.53 | -7.75,12.81 | 0.629 |  |
| Sat after vs before | -14.97 | -31.50,1.56 | 0.076 |  | 3.67 | -7.60,14.94 | 0.523 |  |
| *Other* | | | | | *Other* | | | |
| Mon after vs before | 0.28 | -35.01,35.57 | 0.988 |  | -7.25 | -31.95,17.44 | 0.564 |  |
| Tues after vs before | -14.95 | -42.68,12.78 | 0.290 |  | 9.60 | -13.17,32.36 | 0.408 |  |
| Wed after vs before | -0.47 | -29.34,28.39 | 0.974 |  | 11.51 | -14.77,37.80 | 0.390 |  |
| Thurs after vs before | -9.93 | -39.22,19.36 | 0.505 |  | -21.54 | -47.95,4.87 | 0.110 |  |
| Fri after vs before | 11.06 | -24.97,47.08 | 0.546 |  | -3.84 | -32.05,24.38 | 0.789 |  |
| Sat after vs before | -34.62 | -72.82,3.58 | 0.076 |  | 5.69 | -20.39,31.77 | 0.669 |  |

*Cochran’s Q test to test for heterogeneity between strata. Results presented by first strata of each sociodemographic characteristic.
